# Gamma-hydroxybutyrate to promote slow-wave sleep in major depressive disorder: a randomized crossover trial

**DOI:** 10.1101/2024.10.02.24314769

**Authors:** Francesco Bavato, Laura K. Schnider, Dario A. Dornbierer, Julia R. Di Floriano, Nicole Friedli, Marina Janki, Boris B. Quednow, Hans-Peter Landolt, Oliver G. Bosch, Erich Seifritz

**Author notes:** **Corresponding author:** Francesco Bavato, MD, Department of Adult Psychiatry and Psychotherapy, University Hospital of Psychiatry Zurich, University of Zurich, Lenggstrasse 31, CH-8032 Zurich, Switzerland.

## Abstract

In major depressive disorder (MDD), main clinical features include insomnia and increased daytime sleepiness. However, specific treatment options to promote sleep in MDD are limited. Gamma-hydroxybutyrate (GHB, clinically administered as sodium oxybate) is a GHB/GABA_B_ receptor agonist used clinically in narcolepsy, where it promotes restorative slow-wave sleep (SWS) while reducing next-day sleepiness. Therefore, we performed a randomized, placebo- and active comparator-controlled, double-blind, crossover trial to investigate the sleep-promoting properties of GHB in individuals with MDD. Outpatients aged 20-65 years fulfilling the DSM-V criteria for MDD were enrolled. A single dose of GHB (50mg/kg) was compared with a single dose of the clinical competitor trazodone (1.5 mg/kg) and placebo. Of 29 randomized patients, 23 received at least one intervention and were included in the analysis. Primary outcomes were slow wave sleep ([SWS], as % of total sleep time [TST]) assessed by polysomnography and next-day vigilance (median response time and number of lapses in the psychomotor vigilance test [PVT]). GHB robustly prolonged SWS compared to both trazodone and placebo. GHB also prolonged TST and enhanced sleep efficiency (TST % of time-in-bed), while reducing sleep stages N1, N2, and wake-after-sleep-onset. While the median response time on the PVT was unaffected, GHB reduced the number of lapses compared to trazodone and placebo. No serious adverse events occurred. A single nocturnal dose of GHB effectively promotes SWS and shows more favorable effects on next-day vigilance than trazodone and placebo. Future studies should investigate GHB in clinical settings, including repeated administration.

## Introduction

Major depressive disorder (MDD) is a serious mental health condition, which is considered the largest single contributor to global disability according to the World Health Organization (1). Across the clinical presentations of MDD, impairments of sleep are some of the most consistently observed symptoms (2). Sleep disturbances in MDD include prolonged sleep latency, frequent nocturnal awakenings, non-restorative sleep, and daytime sleepiness (3). Polysomnographic (PSG) investigations demonstrated that decreased slow-wave-sleep (SWS) and increased rapid-eye-movement (REM) sleep duration are hallmarks of nocturnal sleep alterations in MDD, although large inter-individual variability is observed (4). Notwithstanding, current pharmacological options to treat sleep in patients with MDD show unsatisfactory outcomes, as most compounds may even reduce restorative SWS and increase daytime sleepiness (5, 6). Gamma-hydroxybutyrate (GHB, clinically administered as sodium oxybate) acts as agonist at GABA_B_ and GHB binding sites, which modulate homeostatic functions such as eating, sexual behavior, and sleep (7-9). The latter effect is clinically used in narcolepsy, where GHB strongly enhances nocturnal SWS, while reducing next-day sleepiness and cataplexy (10). Potential clinical applications of GHB to treat excessive daytime sleepiness have been suggested in different neuropsychiatric disorders including fibromyalgia and Parkinson’s disease (11, 12). However, to our knowledge, no study has specifically investigated nocturnal GHB administration in MDD to date.

Based on these considerations, we conducted a randomized, balanced, double-blind, placebo-controlled, crossover trial to investigate the effects of a single nocturnal administration of GHB vs. trazodone vs. placebo in patients with MDD. Our primary objectives were to determine the effect of GHB on SWS and next-day sustained attention (assessed with the psychomotor vigilance test [PVT]). Secondary objectives included self-reported sleep quality, sleepiness post-awakening, and next-day affective state. The single nocturnal administration of GHB aimed at testing the efficacy of single add-on treatments on sleep and next-day mental state. This approach might increase the feasibility of GHB administration in various clinical settings, not requiring titration, prolonged administration, or follow-up monitoring (13). Trazodone, one of the most frequently, off-label prescribed substances to treat sleep disturbances in MDD, was selected because of comparable SWS-promoting effects and relatively short half-life (14). We hypothesized, that both GHB and trazodone would increase the duration of restorative SWS (hypothesis: GHB>trazodone>placebo). In contrast to trazodone, however, we expected GHB to promote next-day sustained attention as measured by performance on the PVT (hypothesis: GHB>placebo>trazodone). This project represents the first proof-of-concept study on GHB as a sleep medication in MDD.

## Methods

### Overview

The study was performed as single-center trial between August 2020 and April 2022 in the sleep laboratory at the Institute of Pharmacology and Toxicology, University of Zurich, Switzerland. Formal approval was obtained by the Swiss Agency for Therapeutic Products (Swissmedic) and the Ethics Committee of the Canton of Zurich (Identifier: 2018-01293). The study was preregistered at ClinicalTrials.gov: NCT04082806. All participants provided written informed consent according to the declaration of Helsinki and received a monetary compensation for their study participation. The study followed a randomized, placebo-controlled, order-balanced, double-blind, crossover design and tested GHB against the clinical competitor trazodone, and placebo (Figure 1). The study protocol consisted of five non-consecutive nights: a screening night including a medical visit and all-night PSG examination, an adaptation night to allow for habituation to the laboratory setting, and three experimental nights (GHB, trazodone, and placebo conditions in random order). The three experimental nights were separated by washout phases of 7 days each. Apart from the results presented here, drug effects on memory consolidation, toxicological urine markers, and neurodegenerative/inflammatory blood markers were also assessed and will be published separately.

**Figure 1.**
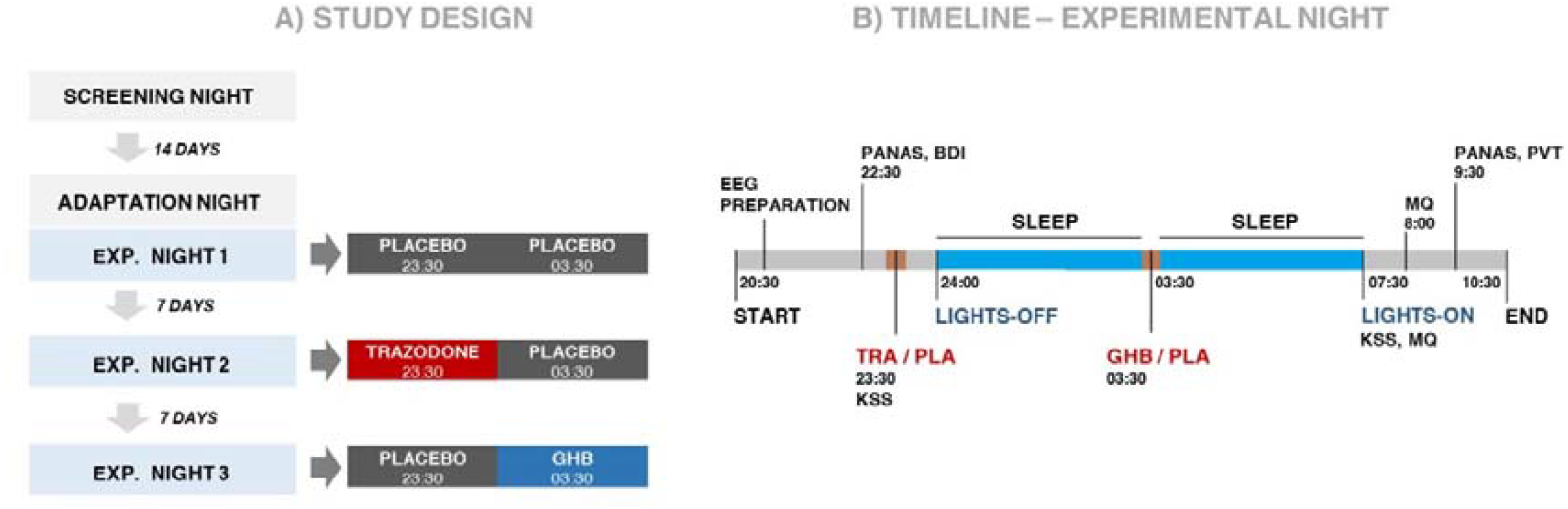
Study design. The study medications were administered at two points during the night, with trazodone or solid placebo at 23:30 and GHB or liquid placebo at 03:30. Abbreviations: PANAS: Positive and Negative Affective Scale; BDI: Beck Depression Inventory; MQ: Morning Questionnaire; PVT: Psychomotor Vigilance Test; KSS: Karolinska Sleepiness Scale; EQ: Evening Questionnaire.

### Participants

Out of 31 patients with MDD initially assessed for eligibility, 29 patients were enrolled into the study and allocated to randomization, 23 received at least one intervention and were included in the analysis, and 22 patients completed the entire study protocol (see CONSORT flowchart, Figure 2). A group of healthy volunteers was also studied to investigate the role of pharmacological sleep enhancement on memory functions but not considered for the present analysis. All participants were assessed by an experienced study physician regarding their general health and psychiatric history. Clinician administered questionnaires included the Montgomery-Åsberg Depression Rating Scale and the Hamilton Depression Rating Scale (HAMD). Self-reported sleep quality was assessed at the screening session with the Pittsburgh Sleep Quality Index. At the beginning of each session, self-reported depressive symptoms were assessed with the Beck Depression Inventory. Inclusion criteria were as follows: diagnosis of MDD according to the Diagnostic and Statistical Manual of Mental Disorders (DSM-5); stable antidepressant treatment (e.g., selective serotonin reuptake inhibitors [SSRI] or serotonin–norepinephrine reuptake inhibitors [SNRI]); age between 20-65 years; no or low dependence on nicotine according to Fagerström Test for Nicotine Dependence (total score <3). Exclusion criteria entailed: intake of any sleep-promoting medication, such as benzodiazepines or z-drugs three days before an experimental night or on a regular basis; any axis-I DSM-5 psychiatric disorder other than MDD; neurological disorders or head injury; any clinically relevant medical diseases. Detailed exclusion criteria are reported in the supplementary methods.

**Figure 2.**
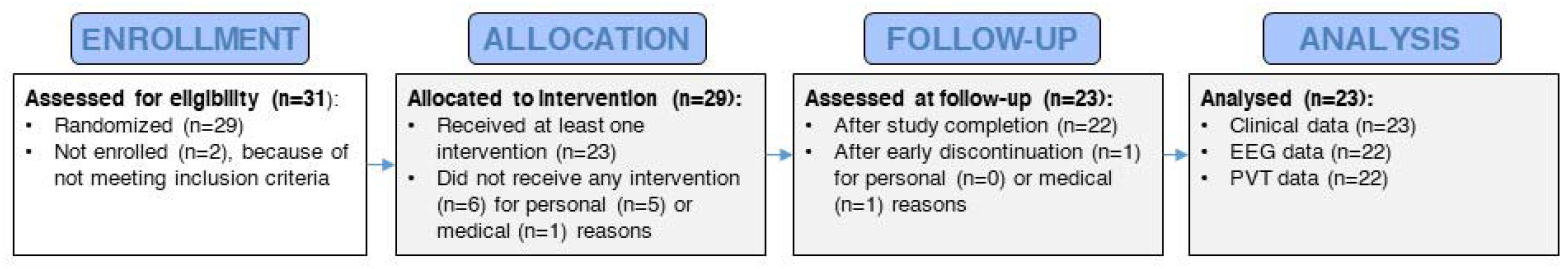
Study protocol diagram according to the CONSORT guidelines. Out of 31 patients assessed for eligibility, 29 patients were enrolled, 23 received at least one intervention and were considered for data analysis, and 22 patients completed the study. Six patients discontinued the study after enrollment but before the first experimental night because of personal (i.e., feeling anxious/unwell in the sleep lab environment, n=2; study protocol being too time consuming, n=2) or medical reasons (i.e., occurrence of new medical conditions, n=2). No participant withdrew from the study because of adverse events. One participant reported use of amphetamines and consequential clinical worsening a few days after the first experimental night (trazodone) and decided to withdraw from the study.

### Interventions

Study medications were administered at two predefined times during each experimental night (Figure 1). Liquid GHB or placebo (0.9% natrium chloride solution) were mixed with 100ml of orange juice (to mask GHB’s salty taste) and given orally at 03:30 after waking up the participants with an acoustic signal without completely turning on the light and while staying in bed. Afterwards, participants were allowed to immediately return to sleep. The median time required for the administration of the liquid drug (from lights-on at 03:30 to lights-off) was 3min (range: 1 to 10min), including an optional toilet break if requested. GHB administration in the middle of the night was chosen because of GHB’s short half-life and the potency to enhance SWS in the second half of the night, when homeostatic sleep pressure is reduced and physiological sleep is typically superficial (13). GHB was dosed according to body weight at 50mg/kg (range 2400-5200mg), based on previous investigations showing a favorable sleep-promoting profile of single administration of GHB at this dose (13).

Trazodone HCl or solid placebo (mannitol) were administered orally at 23:30 and consisted of two capsules matched in appearance and taste. Trazodone was dosed according to body weight at 1.5 mg/kg (range 72-156mg), which is a usual dose for the hypnotic use of the drug, while the antidepressant dose lies at about 300 mg/day (14). Trazodone was administered 30min prior to bedtime, to allow for onset of its sleep-promoting effects (peak concentrations expected 1h after administration) (14).

All study medications were provided by the pharmacy of the University Hospital of Psychiatry Zurich. Balanced, block-wise randomization was performed by an experienced pharmacist at the hospital pharmacy. Both solid and liquid placebos were matched to the appearance and taste of the respective medications to ensure double-blinding of the entire study team.

### Assessments

During experimental nights, PSG was recorded with dedicated amplifiers (SIENNA Ultimate, EMS Handels GmbH) according to the rules of the American Academy of Sleep Medicine (AASM). Recordings consisted of electroencephalography (23 electrodes attached according to the international 10-20 system), bipolar electrooculogram, electromyogram, and electrocardiogram (see detailed description in supplementary methods). Visual sleep stage scoring observing the criteria of the AASM was done by two independent scorers using Rembrandt Analysis Manager (version 8, Embla Systems). Apparent discrepancies between scores were resolved by a third scoring expert. The following sleep variables were computed: (i) total sleep time (TST); (ii) sleep onset latency (SOL); (iii) wake-after-sleep-onset (WASO); (iv) duration of sleep stages (i.e., non-REM [NREM] stage 1 [N1], NREM stage 2 [N2], NREM stage 3 [N3, SWS], and REM sleep); and (v) sleep efficiency ([SE]=□[TST/time-in-bed]□×□100). The duration of sleep stages is reported as percentage of TST.

Each participant’s morning sustained attention was assessed at 09:30 using a 10-min PVT. Reaction times (RT) below 100ms were excluded as false starts. The median RT and the numbers of lapses (trials with RT□>□500 ms) across the 10min were analyzed as indications of sustained vigilant attention (15).

The Positive and Negative Affective Scale (PANAS) was used to assess momentary positive and negative affects before (22:30) and in the morning after (09:30) each experimental night (24:00-07:30). The Karolinska Sleepiness Scale (KSS) was used for the assessment of sleepiness directly before solid drug administration (23:30) and upon awakening (at 07.35). After awakening, a morning questionnaire (MQ, 08:00) was used to assess self-reported sleep quality at each experimental night.

### Outcomes

The primary outcomes were the duration of SWS (% of TST) and the performance on the PVT (median RT and lapses). Secondary were self-reported sleep quality (MQ), post-awakening sleepiness (KSS), and next-day affective state (PANAS). Exploratory outcomes included TST, WASO, and SOL, sleep stages N1, N2, REM, and SE.

### Statistical analysis

All statistical analyses were computed with R-Studio (version 2023.9.1.494, R-Studio Core Team), and JASP. The significance level was set at p<0.05 (two-tailed). Normal *Q*–*Q* and Tukey-Anscombe (residuals vs. fitted) plots were examined to assess model assumptions and goodness-of-fit. Variables with non-normal distributions were log- or power-transformed. To screen for drug effects on primary, secondary and exploratory outcomes, linear mixed-effects models (LMER) for continuous data and generalized linear mixed-effects models (GLM) of the Poisson family for count data were employed (R-package “lme4,” Version 1.1.34). Drug and experimental night order were included as within-subject factor, and subject ID as a random effect. Following model fitting, analyses of variance (ANOVA) were performed, to assess the overall significance of the fixed effects. If the condition was identified as significantly influencing an outcome variable, pairwise comparisons of estimated marginal means for each experimental condition were conducted using the “emmeans” package (Version 1.8.9). Standardized effect sizes (ES) are reported for post-hoc pairwise comparisons using the “eff_size” function of the “emmeans” package. Values of −0.8<ES>0.8 are considered to reflect large effects irrespective of the corresponding p-value. The initial sample size was selected according to power analysis showing that, given a power 1-β of 85%, α=5%, and n=30, medium effects (f=0.21 equivalent to ES=0.42) can be reliably detected (F-test for repeated measure ANOVA, within-subject effects, correlation among repeated measures=0.65).

## Results

### Demographic and clinical characteristics

Sample characteristics are summarized in Table 1. All participants were under stable antidepressant medication and had a current depressive episode with severity level from mild to severe according to HAMD (score≥8). The mean PSQI total score at baseline was above threshold (>5) for poor sleep quality (16).

**Table 1.**
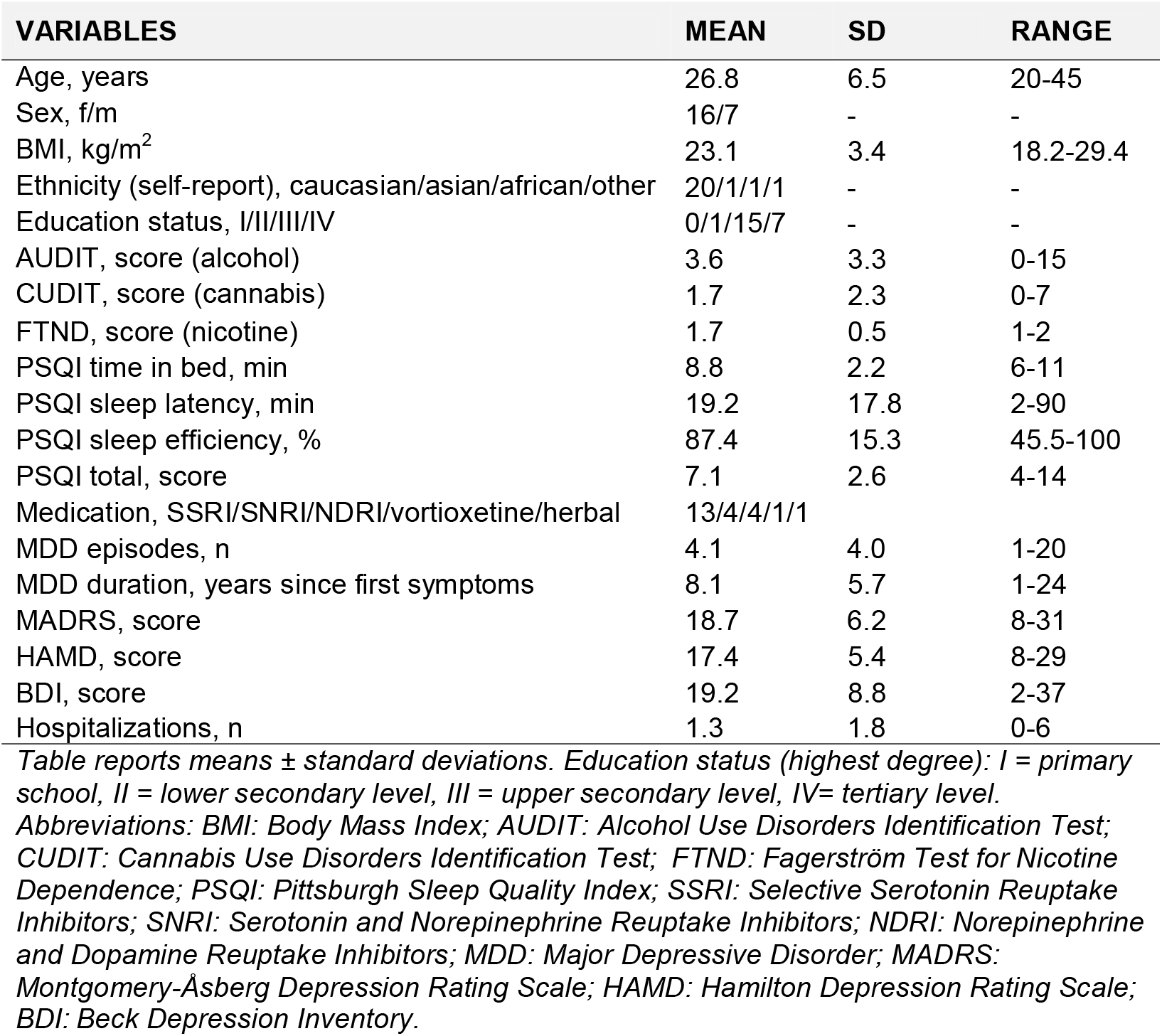
Sociodemographic and clinical characteristics at screening.

### Drug effects on sleep architecture: primary and explorative outcomes

Drug effects on PSG variables are summarized in Figure 3. Compared to placebo, GHB induced a strong elevation of SWS (% of TST: +15.8; 95% CI, +10.8, +20.8 p=<0.001; ES=2.03), while shortening N1 (% of TST: -9.0; 95% CI, -4.6, -1.4; p<0.001; ES=2.01), N2 sleep (% of TST: - 8.5; 95% CI, -13.9, -3.2; p=0.003; ES=1.03) and WASO (minutes: -19.5; 95% CI, -35.5, -3.6; p=0.025; ES=0.74). GHB also increased SE (% of TST: +5.5; 95% CI, +1.1, +9.9; p=0.011; ES=0.84) and TST (minutes: +24.0; 95% CI, +4.5, +43.4; p=0.011; ES=0.85) compared to placebo. Trazodone did not induce a significant prolongation of SWS (% of TST: +3.6; 95% CI, -1.4, +8.6; p=0.151; ES=0.46), but tended to elevate TST (minutes: +18.2; 95% CI, -1.1, +37.6; p=0.087; ES=0.56) and SE (% of TST: +4.3; 95% CI, -0.1, +8.7; p=0.072; ES=0.58), while reducing WASO (minutes: -21.5; 95% CI, -37.4, -5.6; p<0.001; ES=1.26). Compared to trazodone, GHB increased SWS (% of TST: +12.2; 95% CI, +7.3, +17.1; p<.001; ES=1.57), while reducing N1 (% of TST: -4.5; 95% CI, -6.1, -2.9; p<0.001; ES=2.06) and N2 (% of TST: -6.3; 95% CI, -11.6, -1.0; p=0.020; ES=0.75) sleep. No differences between conditions were observed for SOL and the duration of REM sleep.

**Figure 3.**
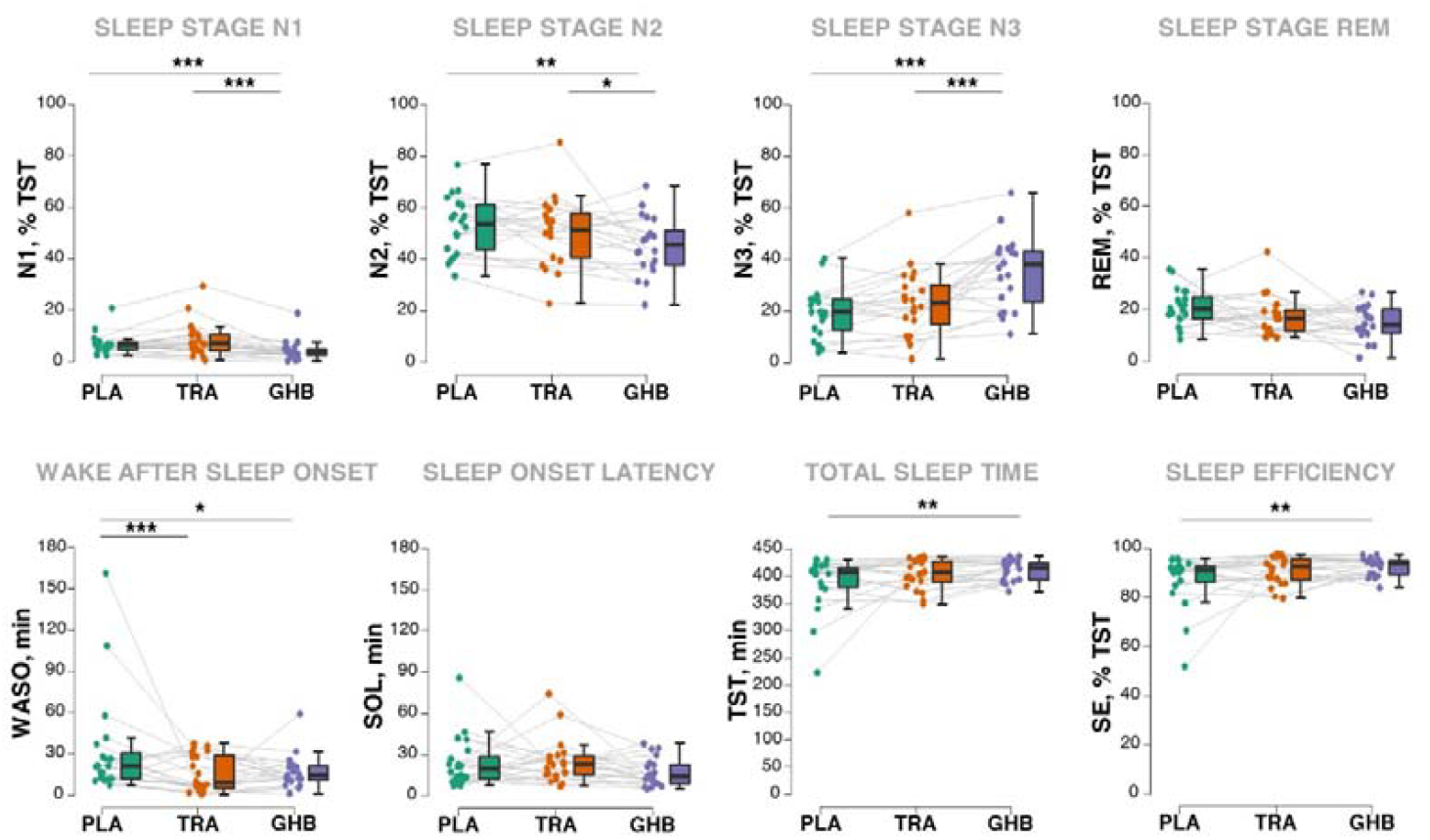
Drug effects on nocturnal sleep architecture. Boxplots and jittered raw data are visualized. The duration of the sleep stages N1, N2, slow-wave-sleep (SWS, N3) and rapid eye movement (REM) sleep are reported as % of total sleep time (TST). The duration of wake-after-sleep-onset (WASO), sleep onset latency (SOL) and TST are reported in minutes. Sleep efficiency (SE) is calculated as SE = (TST) / (total time-in-bed)L×L100.

### Drug effects on next-day vigilance: primary outcome

The statistical analyses of the PVT revealed no drug effects on median RT (LMER with Gaussian distribution) when corrected for experimental night order (Table 2). By contrast, significant drug effects were observed on the number of lapses (GLM with Poisson distribution), including the correction for experimental night order. The post-hoc analyses showed that after GHB, patients produced less lapses than after trazodone (ratio of means: -0.50; 95% CI, -0.21, -0.78; p<0.001; ES=0.50) and placebo (ratio of means: -0.51; 95% CI, -0.21, -0.80; p<0.001; ES=0.51). No difference was observed between the trazodone and placebo conditions (+0.36; 95% CI, - 2.2, +2.9; p=0.95; ES<0.01).

**Table 2.** Drug effects on sustained attention, self-reported sleep quality, post-awakening sleepiness, and affective state.

| VARIABLES | PLACEBO<br>MEAN ± SD | PLA-TRA<br>P-VALUE | TRAZODONE<br>MEAN ± SD | TRA-GHB<br>P-VALUE | GHB<br>MEAN ± SD | PLA-GHB<br>P-VALUE | P-VALUE<br>CONDITION |
| --- | --- | --- | --- | --- | --- | --- | --- |
| PVT median RT | 307.8 ± 44.2 |  | 320.3±44.7 |  | 307.5±33.8 |  | 0.16 |
| PVT lapses | 5.5 ± 7.6 | 0.95 | 7.7±11.8 | **<0.001** | 3.4±3.6 | **<0.001** | **<0.001** |
| MQ awakening, n | 2.1 ± 1.3 |  | 1.7 ± 1.4 |  | 1.6±1.3 |  | 0.28 |
| MQ time awake, min | 26.4 ± 50.5 |  | 18.8 ± 29.8 |  | 12.7±15.7 |  | 0.06 |
| MQ superficial sleep, VAS | 43.6 ± 16.1 |  | 45.8 ± 22.4 |  | 38.3±18.7 |  | 0.38 |
| MQ undisturbed sleep, VAS | 48.8 ± 16.1 |  | 55.9 ± 18.7 |  | 53.0±20.1 |  | 0.30 |
| MQ awakening recovery, VAS | 38.6 ± 18.2 | 0.059 | 29.2 ± 20.3 | **0.019** | 41.8±25.8 | 0.64 | **0.043** |
| MQ awakening energy, VAS | 45.9 ± 12.5 | 0.086 | 39.6 ± 15.9 | **0.003** | 51.4±17.0 | 0.20 | **0.012** |
| KSS pre-sleep, score | 6.2 ± 2.1 |  | 5.8 ± 1.8 |  | 5.6±1.9 |  | 0.26 |
| KSS post-awakening, score | 6.1 ± 2.1 | **0.028** | 7.3 ± 1.6 | **0.014** | 5.9±2.4 | 0.81 | **0.025** |
| PANAS positive evening, score | 22.3 ± 5.2 |  | 22.3 ± 6.0 |  | 21.8±6.6 |  | 0.85 |
| PANAS negative evening, score | 16.0 ± 5.4 |  | 16.6 ± 5.7 |  | 15.1±4.1 |  | 0.35 |
| PANAS positive morning, score | 24.0 ± 7.8 |  | 20.7 ± 8.0 |  | 23.5±7.5 |  | 0.15 |
| PANAS negative morning, score | 14.5 ± 4.7 |  | 15.0 ± 3.9 |  | 14.6±3.3 |  | 0.92 |

*Table reports means ± standard deviations. Model statistics refers to LMERS with condition (placebo vs. trazodone vs. GHB) and night order as factors. Significance levels of posthoc pairwise comparisons between conditions are reported if the model was found significant for the variable condition. Abbreviations: PVT: Psychomotor Vigilance Test; RT: response time; MQ: Morning Questionnaire; VAS: Visual Analogue Scale (1-100); KSS: Karolinska Sleepiness Scale; PANAS: Positive and Negative Affective Scale.*

### Drug effects on self-reported sleep quality, post-awakening sleepiness, next-day affective state: secondary outcomes

Results of self-reported sleep quality (MQ), post-awakening sleepiness (KSS), and affective state (PANAS) across conditions are summarized in Table 2. Trazodone increased sleepiness upon awakening (KSS) compared to GHB (+1.4; 95% CI, +0.30, +2.5; p=0.014; ES=0.53) and placebo (+1.3; 95% CI, +0.15, +2.9; p=0.028; ES=0.69) conditions and reduced next-day energy level (−12.5; 95% CI, - 20.5, -4.4; p=0.003; ES=0.94) and recovery feeling (−13.1; 95% CI, -23.9, -2.3; p=0.019; ES=0.74) compared to GHB. No drug effects on affective state (PANAS) were observed.

### Safety and tolerability

No serious adverse events occurred. In the GHB condition, 8 of 22 participants reported mild side effects, with nausea and dizziness being the most common. In the trazodone condition, 1 of 23 participants reported moderate and 7 of 23 mild side effects, with nausea and headache being the most common. No side effects were observed in the placebo group (0 of 22). No participant withdrew from the study because of adverse events. Notably, a predominance of side effects in female participants was observed for GHB (female vs. male, n=8 of 15 vs. 0 of 7) but not for trazodone (female vs. male, n=6 of 16 vs. 2 of 7). In the overall sample, there was no emergence of serious suicidal ideation as assessed by the suicidality screening form.

## Discussion

We demonstrated that GHB strongly increases SWS and reduces next morning attentional lapses in patients with MDD. GHB showed a more favorable sleep-promoting profile compared to trazodone, which currently represents one of the most often administered substances to treat disturbed sleep in MDD.

The observed effects of a single nocturnal administration of GHB on SWS are in line with our previous observation in healthy volunteers and have relevant clinical implications (13). Deep SWS has crucial roles in physiological brain processes (e.g., cellular homeostasis, synaptic plasticity, and removal of waste metabolites) and high-order brain functions (e.g., cognitive performance, memory consolidation, and affective state) (17). The reduction of SWS in many MDD patients has been linked to affective symptoms, cognitive deficits, and neurodegenerative alterations (18). In contrast, increased SWS has been associated with a reduction of depressive symptoms in naturalistic and experimental studies of MDD (19-21). Therefore, SWS is considered a main clinical target to develop treatment interventions for MDD.

Among the current pharmacological options for the treatment of sleep disorders in psychiatry, most compounds either reduce SWS (e.g., benzodiazepines and z-drugs) (5) or increase daytime sleepiness (e.g., sleep-promoting antidepressants and second-generation antipsychotics) (22). Thus, the observation of increased SWS while improving next-day vigilance represents a unique advantage of GHB over other current sleep medications. In the current study, GHB was associated with a reduction in PVT lapses, which are thought to be the result of perceptual, processing, or executive failures associated with reduced cognitive alertness. An increase in PVT lapses has been consistently associated with sleep deprived states and fatigue (23). Thus, our results confirm that nocturnal GHB administration might promote next-day vigilance even after a single administration (24).

Despite inducing a strong prolongation of SWS and promoting next-day vigilance, GHB was not associated with an improvement of subjective sleep quality or next-day affective state. A similar discordance between subjective and objective measures of sleep have been frequently reported in people with insomnia (25). Nonetheless, in consideration of the suggested causal link between SWS and depression, an improvement of affective state after SWS-promotion by GHB might have been expected. In previous neuroimaging studies, we demonstrated that a single nocturnal administration of GHB results in next-day neural adaptations (i.e., of functional brain networks and of glutamate metabolism), which could hint at rapid antidepressant properties (24, 26). However, investigations on GHB in narcolepsy, Parkinson’s disease, and fibromyalgia suggest that repeated administration of GHB over 4-6 weeks are required to elicit favorable effects on symptom dimensions such as fatigue, cataplexy, and pain sensitivity (10-12). Sleep-promoting antidepressants like trazodone and mirtazapine also require several days of treatment to show antidepressant properties (14). Future investigations including repeated administrations are therefore warranted to clarify potential effects of GHB on subjective sleep quality, affective state and depressive symptoms. Moreover, the recent development of extended-release GHB formulations, which prolong the sleep-promoting effects of GHB over the entire night, might be particularly indicated for future studies (27).

Our study bears several limitations. The final sample size of n=23 resulted in lower statistical power than initially planned, thus, affecting the generalizability of our findings, especially regarding exploratory outcome variables. Nonetheless, the large effect size of the main outcome variable (ES for SWS=2.03, n=23, α=5%) still allowed sufficient power (1-β=0.99). The current investigation did not provide information on individual characteristics predicting treatment responses also because of insufficient sample size to perform subgroup analysis. Even though GHB was overall well-tolerated, the higher prevalence of side effects in female participants might limit the applicability of GHB in a clinical context and clearly requires further investigations. Sex-dependent adaptations of the dose (lower dose or slower titration) might be necessary although larger studies in narcolepsy and fibromyalgia did not report similar sex-dependent differences in the tolerability.

It is important to note, that GHB can induce pharmacological tolerance, and its regular use may carry the risk of developing a substance use disorder (28). However, the addictive potential of GHB (and its related effects on midbrain dopamine circuits) varies greatly with dose, with repeated low doses (recreational/non-medical use) paradoxically showing much higher addictive potential (and dopamine release) than higher doses (medical use) (29). Coherently, the medical use of GHB in patients with narcolepsy with cataplexy is associated with a very low risk of addiction (30). In any case, a cautious consideration of the risks and benefits of GHB use in psychiatry should await future evidence from trials conducted in the clinical context.

In conclusion, we provide evidence of favorable sleep-promoting effects of GHB in patients with MDD. This proof-of-concept study represents the first experimental validation of single GHB administration to promote sleep in a clinical psychiatric population. Overall, our findings support further investigation of GHB as a promising sleep medication candidate in patients with MDD and in psychiatry in general.

## Funding & Acknowledgements

The study was supported by a grant from the Swiss National Science Foundation (SNSF) to ES (grant # 175780). FB received personal financial support from SNSF (grant # 217637).

All authors report no conflicts of interest, financial or otherwise. We thank Anna Habereder, Rahel Gelin, Thomas Monasterios Gallardo, Viveka Boller, and Corinne Eicher for the support with data collection, Helena Jenzer for the support with study planning, and Benjamin Stucki for the support with statistical analysis.

## Data Availability

All data produced in the present study are available upon reasonable request to the authors

## Notes

### Competing Interest Statement

The authors have declared no competing interest.

### Clinical Trial

NCT04082806

### Author Declarations

Formal approval was obtained by the Swiss Agency for Therapeutic Products (Swissmedic) and the Ethics Committee of the Canton of Zurich (Identifier: 2018-01293)

